# The curious case of lower reported racial discrimination in healthcare

**DOI:** 10.64898/2026.02.27.26347279

**Authors:** Didier Ruedin, Denise Efionayi-Mäder, Irina Radu, Alessandra Polidori, Lisa Stalder

## Abstract

**Objective:** Explore self-reported racial discrimination in healthcare.

**Methods:** Representative population sample, Switzerland, repeated cross-sectional data 2016 to 2024 (N=15,525).

**Results:** Contrary to expectation, respondents from the migration-related population (foreign citizens, foreign born, migration background, first/second generation) report less racial discrimination than members of the majority population. Over time, we see an increase in the non migration-related population reporting (racial) discrimination in healthcare, while the share for the migration-related population is constant. The validity of the instrument is demonstrated with reported discrimination at work and in housing and the results are reliable across specifications and statistical controls.

**Conclusion:** We speculate that in some cases, reported racial discrimination may express unmet expectations in healthcare more generally.

## Introduction

The experience of discrimination is associated with lower health outcomes across different indicators of physical and mental health (Emmer, Dorn, and Mata 2024; Vines et al. 2017). Existing evidence often draws on broad questions of the self-reported experience of discrimination, such as “Have you experienced prejudice or discrimination in the last 24 months?”, differentiating reasons such as gender identity, sex, disability, or ethnicity. To date, details such as the situation in which discrimination is experienced are not systematically assessed when discrimination and health outcomes are associated. Apart from qualitative evidence on differential treatment in healthcare (Hamed et al. 2022; Zemouri et al. 2024; Wegelin et al. 2021), we have no indicators specific to healthcare.

In this contribution, we use a large-scale representative sample to examine who experiences racial discrimination in healthcare. If qualitative evidence generalizes beyond the specific contexts that are studied, we should find racial discrimination in the population survey. In Western Europe racial discrimination is often linked to perceived migration background, affecting refugees, labour immigrants, and their children (Heath and Schneider 2021; Ball, Steffens, and Niedlich 2022). However, contrary to expectations, we find that migration-related populations are *less* likely to report discrimination in healthcare, irrespective of how they are captured in the data. We can observe an increase in the majority population reporting racial discrimination in healthcare, which theory cannot explain and we interpret as a possible expression of unmet expectations.

## Data and Methods

### Outcome variable: experience of racial discrimination in healthcare

To capture the experience of racial discrimination in healthcare, we rely on a widely used question of self-reported discrimination. Part of a multi-stage question, respondents are first asked: “During the past five years, have you experienced a situation where you have been discriminated against due to your belonging to a group?”. A definition of discrimination was provided as a pop-up: “Discrimination occurs when people are treated unfairly or intolerantly, humiliated, threatened or endangered because they belong to a certain group (e.g. women, Black people, etc.).”. Irrespective of the pop-up, the web version also stated “For example with regard to your nationality, your religion, your sexual orientation, your age, etc.”. The exact wording of these questions was determined after extensive testing in focus groups and cognitive testing.

Subsequently, respondents were asked: “Where did this discrimination happen?” to differentiate between discrimination in Switzerland and abroad (not retained). For discrimination in Switzerland, the motive was captured as: “Due to your belonging to which group or on which ground have you been a victim of discrimination?”, offering 15 non-exclusive motives plus an option to write in other reasons. The list of motives follows similar surveys in other countries and was revised after pilot testing and an evaluation of open responses. For this article, we include the following motives as racial discrimination: nationality/citizenship, ethnicity, religion, skin colour/body features, language/accent. Cognitive testing and an evaluation of a follow-up question during the pilot confirms that respondents mean non-Christian religion and non-local languages. The same categorization was previously used in Germany (Von Dem Knesebeck and Klein 2024). For empirical robustness, we also excluded language and religion (‘narrow sample’), and considered any motive for a broader concept of discrimination more generally (‘any discrimination’). Until 2020, the questionnaire referred explicitly to ‘foreign nationality’, since 2022 the question mentions ‘nationality’. Cognitive testing and an open question during the pilot confirm that respondents think of ‘foreign’ nationality, and we show that the reported changes are unlikely driven by this slight change in wording. We only considered responses of self-reported discrimination in healthcare in the subsequent question: “In which precise situations have you been a victim of discrimination?”. 14 situations are offered, plus an open category.

### Predictor: migration−related population

Our predictor captures whether a respondent can be considered part of the migration-related population, for which we have four indicators. The multiple indicators provide robustness. First, we can identify respondents with foreign citizenship (as opposed to Swiss or double citizens). Second, we can identify respondents who were born abroad. Third and fourth, we can capture migration background according to the Swiss Federal Statistical Office (BFS/OFS). On the one hand as a binary variable (migration background or not), on the other hand differentiating between migration background of the first and migration background of the second generation. According to this definition, the migration background is defined by a combination of country of birth and nationality (current and at birth) of the respondent, as well as the country of birth of the parents (**appendix 1**). All these variables are based on registered characteristics. This means that they do not capture self-reported minority status, nor the respondents’ perception that they are being read or seen by others as ‘foreign’ (compare Fuchs et al. (2025); Borrelli and Ruedin (2024)). In Switzerland the majority of this population is migration-related, so these four approaches should include racial and ethnic minorities.

### Control variables

In the regression models, we control for respondents’ age (grouped into 3 categories: <30, 30 to 49, and 50+), sex (male, female, according to SAGER protocol (Epps et al. 2022)), level of education (compulsory schooling, secondary II, tertiary level), whether they live in an urban place (densely populated, intermediate, thinly populated area), and the time in Switzerland (calculated based on the year of arrival for respondents born abroad; not included in models based on country of birth). As a survey administered by the Swiss Federal Statistical Office, the control variables available are similar to what is available in register data. They were chosen as they may affect the awareness of discrimination, as well as exposure to differential treatment in healthcare.

### Data: Representative survey

The survey *Vivre Ensemble en Suisse* (Survey on diversity and coexistence in Switzerland) is a nationally representative register-based sample of the population aged 15 to 88 years, carried out by the Swiss Federal Statistical Office and the Service for the fight against racism (FRB/SLR), fielded every two years (BFS 2024). Respondents who experience discrimination in healthcare are relatively rare (**appendix 2**), but the representative nature of the data allows us to express the prevalence of discrimination — particularly when looking at changes over time. We include all samples between 2016 and 2024 (N=15,525). For robustness, we consider a narrow definition of racial discrimination and those reporting any kind of discrimination (**appendix 3**).

### Analytical strategy

Analytically, we describe differences in prevalence (as percentages to account for different sample sizes), and use Bayesian regression models with control variables (Martin, Quinn, and Park 2011; Goplerud 2022) in *R* (2025). We use the default non-informative priors that by design reduce the influence of highly influential observations in small (sub-)samples (Kruschke 2015).

## Results: Increasing discrimination by the non migration−related population

In Figure 1 we can see an increase in self-reported discrimination in healthcare, but it is respondents of the *majority* population who report more racial discrimination (panel A). The prevalence of discrimination was similar for the two populations in 2016 and 2018, but since 2020, we observe an increase among the non migration-related population. In **appendix 4** we show that this increase can be observed for all 4 definitions of the migration-related population available in the data. In panel B and panel C, we include the prevalence of self-reported discrimination at work and in housing — situations where discrimination is persistently higher than in healthcare. We can see that the lines for the majority population and for the migration-related population are roughly parallel and that the migration-related population experiences more racial discrimination, as can be expected. This demonstrates that neither the instrument nor the sample are problematic.

**Figure 1:**
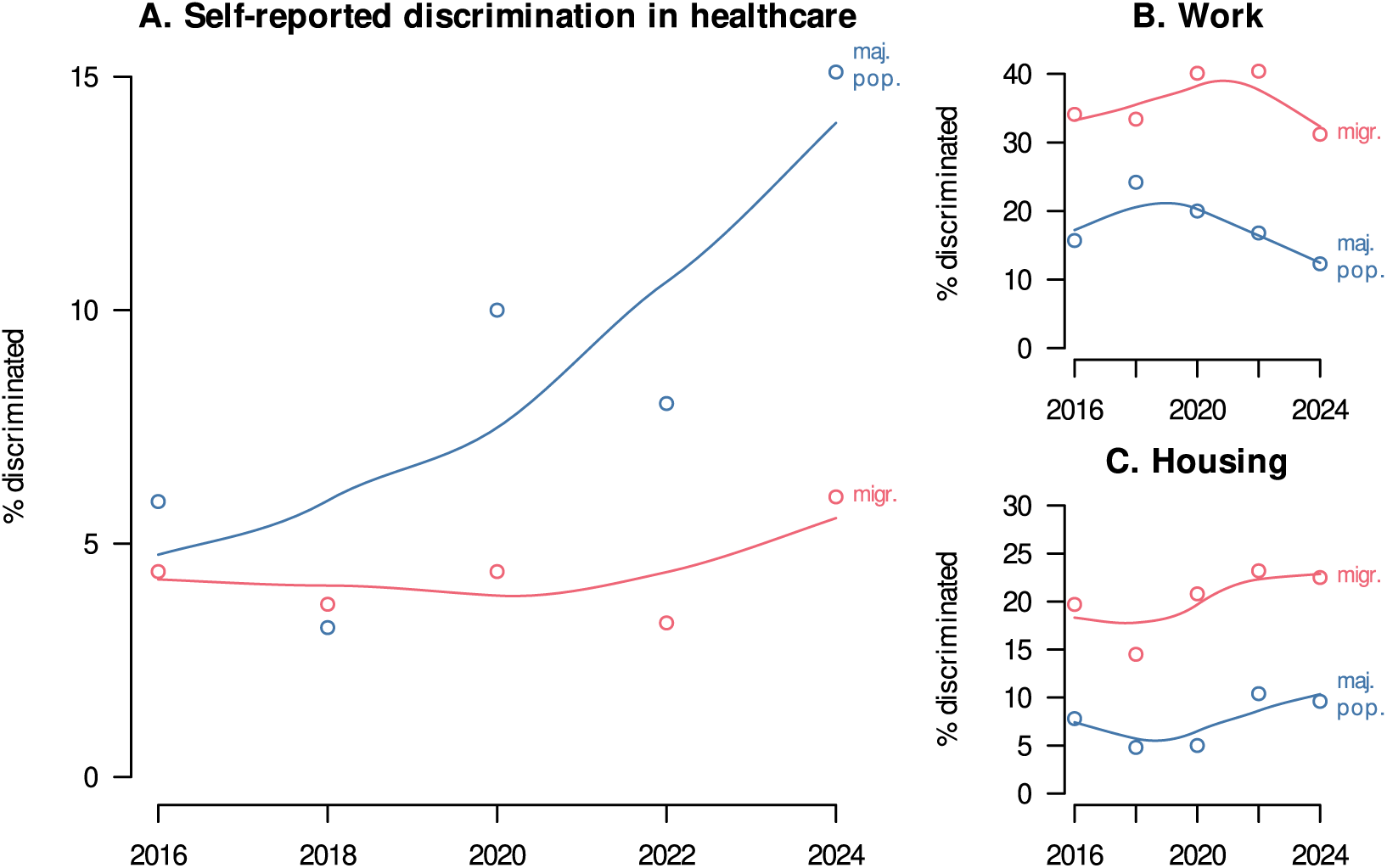
Self-reported experience of racial discrimination in healthcare (panel A), work (panel B), and housing (panel C) by migration background, Switzerland, 2016 to 2024. The circles give the percentages for the majority population (blue) and respondents with a migration background (red). The lines are LOESS smoothed with span=0.9.

In addition to showing equivalent developments when migration-related populations are defined by country of birth, citizenship, or when first- and second-generation migration background is differentiated (**appendix 4**), we can also show this pattern of higher levels of self-reported discrimination in multiple regression models that account for other factors (**appendix 5**). We can see that respondents from migration-related populations consistently report *less* discrimination in healthcare than members of the majority population in 2024. We find the same patterns when all samples are combined and changes over time are accounted for in a variable (**appendix 6**) and when using hierarchical models (**appendix 7**). Pushing the limits of small sample sizes, we look at individual motives in **appendix 4**. While the described pattern can typically be found for individual motives, this is especially the case for self-reported discrimination based on skin colour and language, as well as when using migration background rather than country of birth.

In **appendix 8** we account for the time since respondents from migration-related populations have come to Switzerland. In line with the results for the distinction between migration background of the first generation and migration background of the second generation in **appendix 8**, we find no substantial differences in the experience of discrimination in healthcare. Similarly, in **appendix 9** we find no difference in reports of discrimination in healthcare between the German speaking area of Switzerland and the French and Italian-speaking areas. The inclusion of an interaction effect between the year of the survey and the precise definition of migration-related population in **appendix 10** confirms the robustness of the findings, as does the control for political ideology in **appendix 11**. In **appendix 12** and **appendix 13** we show how patterns of reported discrimination at work and in housing follow the expectations in the literature: higher levels of discrimination for migration-related populations (Flage 2018; Zschirnt and Ruedin 2016). **Appendix 14** replicates stable levels of discrimination in healthcare among a sample of recent immigrants to Switzerland 2016 to 2024.

## Discussion and conclusion

Using a large-scale representative survey, we provided consistent evidence that migration-related population report *less* racial discrimination in healthcare. This contradicts a cross-sectional population survey in Germany in 2022 (Von Dem Knesebeck and Klein 2024) which focused on different forms of discrimination in healthcare — the only such survey we could identify — and qualitative evidence from Switzerland among others which demonstrates that some members of migration-related populations experience discrimination (Wanner and Pecoraro 2023; Duveau et al. 2023; Eriksson, Carlbom, and Essén 2025). With the repeated representative population survey, we should be in a better position to capture the prevalence and developments of discrimination over time than existing work. We find that in the past five years or so, self-reported racial discrimination by the majority population has increased, while it remained relatively stable for the migration-related population.

Robustness checks indicated that the results are not driven by any particular motive of discrimination, but rather a mix of dimensions associated with racial discrimination. Pushing small numbers when disaggregating motives, we find the clearest increase for skin colour and language. The latter precludes the possibility that the observed increase is due to a growing population of people whose grandparents have moved to the country (‘third generation’). However, beyond (not) belonging to a migration-related population, we could not identify variables that are consistently associated with greater experience of racial discrimination in healthcare. In their tendency, more education may be associated with slightly less, older with slightly more, and women possibly slightly more racial discrimination in healthcare (compare Zemouri et al. (2024) who reported gender differences in racial discrimination in the Nether-lands), but these associations are suggestive at best.

The results are clear that the non migration-related population increasingly report discrimination in healthcare on grounds of nationality, skin-colour, language, religion, or ethnic origin. While healthcare is characterized by lower levels of discrimination than say housing or employment (Figure 1, Pattillo et al. (2023)), the high professionalization and the widespread use of checklists and standardized tools cannot explain the changes over time. Similarly, the migration-related population may have lower expectations, something generally explained with a reference frame in country of origin (Röder and Mühlau 2012; Luiking et al. 2019), the experience of discrimination at work or in housing, or attempts not to be perceived as ‘difficult’ patients when calling out racial discrimination (Holmes et al. 2021). These factors may lead to under-reporting discrimination but do not explain the changes over time.

We are left with an unexpected empirical regularity. It is conceivable that the majority population have high expectations of healthcare in Switzerland, and disappointment with service delivery may be expressed as ‘racism’, especially in a survey where questions of racial and ethnic differences are clearly present. In this reading of the evidence, what we capture may be expressions of a perceived ‘decline’ in healthcare, possibly a reaction to doctors and healthcare providers who were not born in Switzerland, who do not speak the local dialect or language fluently or have a different skin colour, or seeing leaflets translated into foreign languages. Indeed, the complex and fragmented nature of the Swiss healthcare system may contribute to the reported racial discrimination by members of the majority population, because if something goes wrong, it can be difficult to figure out why. Vague disappointment with service delivery and possibly a feeling that ‘others’ get more may then expressed in the form of (reverse) ‘racism’ — even though there are no historically rooted power differences presents, so academics would not apply this label (Bonnett 2022; Quijano 2000). These perceptions that ‘others’ get something (and be this just a translation) and ‘me’, I get nothing special may be more visible in healthcare because levels of discrimination are probably lower than in other domains. Recent research in sociology suggests that such unmet expectations may affect as much majority as minority populations (Tubergen 2026; Genoni and Ruedin 2025). A mixed-methods study of complaints in Swedish healthcare linked the experience of ethnic discrimination to subtle forms of discrimination and unmet expectations that according to the authors may be explained by misunderstandings and communication problems (Eriksson, Carlbom, and Essén 2025). The examples they cite refer to ethnic minorities but also note that unmet expectations may be due to pressured care situations that are perceived as disrespectful treatment. With the data available, we cannot examine individual incidences, nor can we support our hunch beyond the observation that the described tendency to report more discrimination can be observed for racial discrimination as well as other forms of discrimination (‘any’ sample).

The possible interpretation of a purported ‘reverse racism’, possibly a backlash, calls for deeper investigation in future studies. We need to find out whether the context of the survey encourages such responses by the majority population, and whether the self-reported racial discrimination by the majority population has the same negative effects on health as it has for the minority population (Emmer, Dorn, and Mata 2024; Vines et al. 2017). While interpersonal racism has a stronger impact on mental health than structural racism (Boma Lazaridou 2025), we encourage researchers to study structural forms of racism (Adkins-Jackson et al. 2021, for suggestions in healthcare), which may be less influenced by individual perceptions. By encouraging researchers to take seriously the pattern we describe here, however, we urge practitioners to continue efforts to reduce and eliminate racism in healthcare, to continue efforts to name and reflect racism, and to continue professionalization (Blandenier et al. 2025). What we suggest is to add considerations how members of the majority population perceive such efforts and communicate their needs and expectations in healthcare provision. It may well come down to better communicating the meaning and implications of equity on which healthcare provision is based.

## Data Availability

The data used are available upon reasonable request from the Swiss Federal Statistical Office (https://www.bfs.admin.ch/bfs/en/home/statistics/population/surveys/zids.html).

## Ethics

Secondary analysis of anonymous data; exempt from ethics review.

## Acknowledgements and Author Contributions

This research was supported by the Swiss National Science Foundation project grant 200939, with additional support from the NCCR on the move (grant 51NF40_205605). We would like to thank Johanna Probst for comments. *Author contributions*: DR (conceptualization, formal analysis, writing - original draft, writing - review & editing), DE (writing - review & editing), IR (writing - review & editing), AP (writing - review & editing), LS (writing - review & editing).

## Appendix

## 1. Defining migration background

Table 1 outlines how migration background is defined according to the Swiss Federal Statistical Office (BFS/OFS). A person can have: no migration background (0), a migration background of the first generation (1), or a migration background of the second generation (2). In the binary definition, a distinction is made between no migration background (0) and a migration background (1 or 2).

**Table 1:**
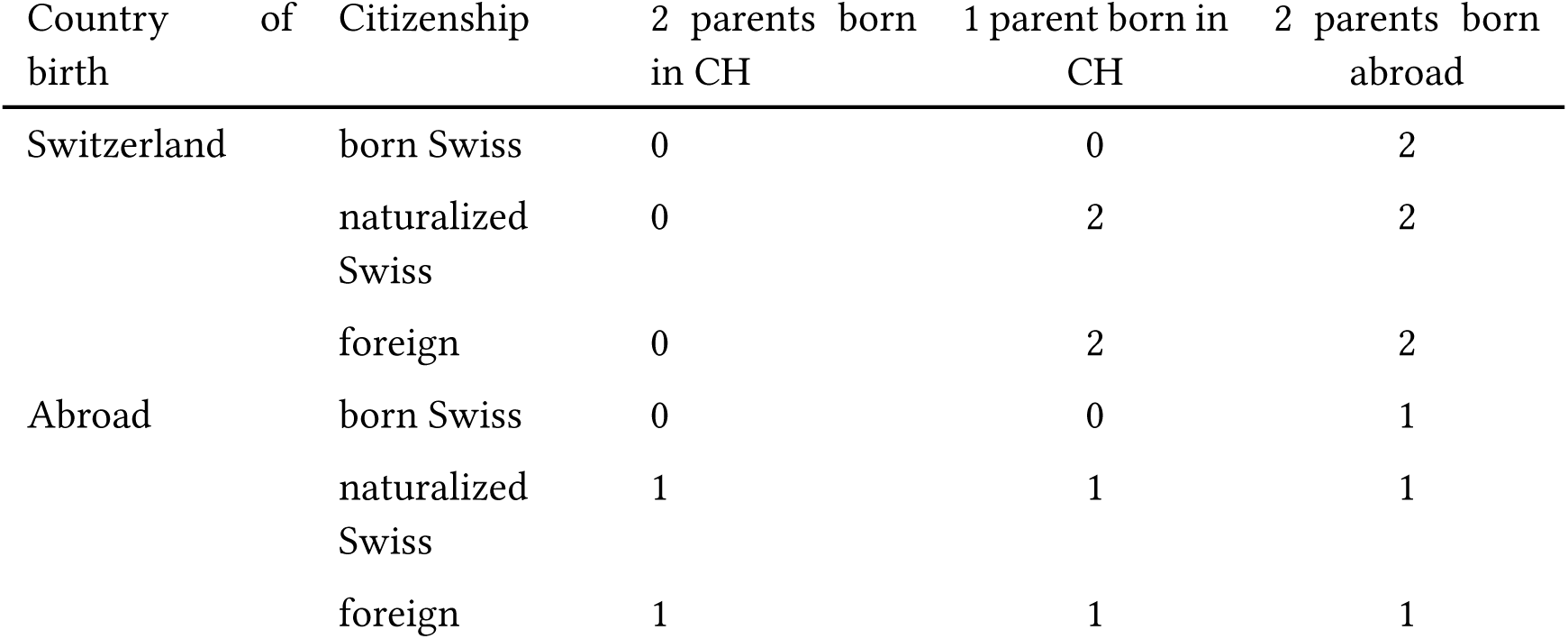
Migration background according to the Swiss Federal Statistical Office (BFS/OFS)

## 2. Motives of discrimination included in the different samples

Table 2 gives an overview which motives of discrimination are included in the different samples.

**Table 2:** Overview of the motives of discrimination included in the different samples.

| Motive | Any | Broad | Narrow |
| --- | --- | --- | --- |
| Age | yes |  |  |
| Sex | yes |  |  |
| Sexual orientation | yes |  |  |
| Nationality (citizenship) | yes | yes | yes |
| Ethnicity | yes | yes | yes |
| Religion | yes | yes |  |
| Skin colour, body features | yes | yes | yes |
| Language, accent | yes | yes |  |
| Socio-economic level | yes |  |  |
| Job | yes |  |  |
| Political opinions, other opinions | yes |  |  |
| Disability | yes |  |  |
| Gender identity | yes |  |  |
| Family situation | yes |  |  |
| Weight | yes |  |  |

## 3. Size of subsamples

**Table 3:** Size of subsamples used in the analysis.

| Population | N in 2016 | N in 2018 | N in 2020 | N in 2022 | N in 2024 |
| --- | --- | --- | --- | --- | --- |
| Total sample | 3,010 | 3,127 | 3,258 | 2,908 | 3,222 |
| Experienced discrimination (any form) | 617 | 689 | 854 | 729 | 802 |
| Experienced racial discrimination (broad, any domain) | 281 | 368 | 432 | 428 | 449 |
| Experienced racial discrimination (narrow, any domain) | 237 | 249 | 266 | 263 | 239 |
| Experienced racial discrimination in healthcare | 29 | 26 | 53 | 53 | 82 |

## 4. Increase of self−reported discrimination for all definitions and sub− samples

In this section we show changes in reported discrimination in healthcare over time, using a different definition of the migration-related population each time. We can observe substantively equivalent developments, suggesting robust findings.

### Migration background (2 categories)

In Figure 2, migration-related is captured as migration background (2 categories). The figure shows that the increase in reported discrimination can be found in all samples, while it is least pronounced when ‘nationality’ is the only motive of discrimination.

**Figure 2:**
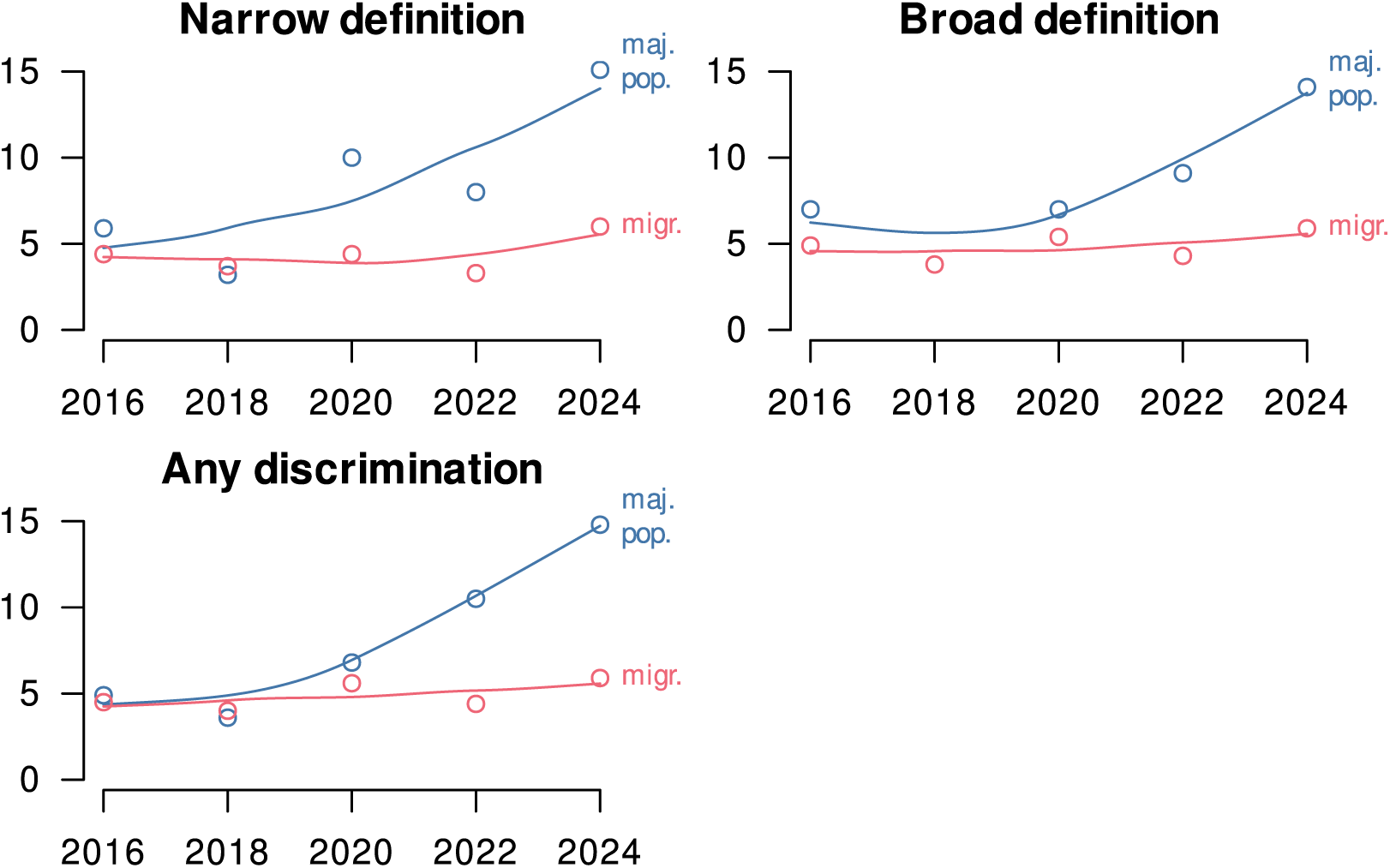
Difference in reported discrimination in healthcare by migration background (2 categories), in percentage points, Switzerland, 2016 to 2024. Different definitions of discrimination are used in each panel.

**Figure 3:**
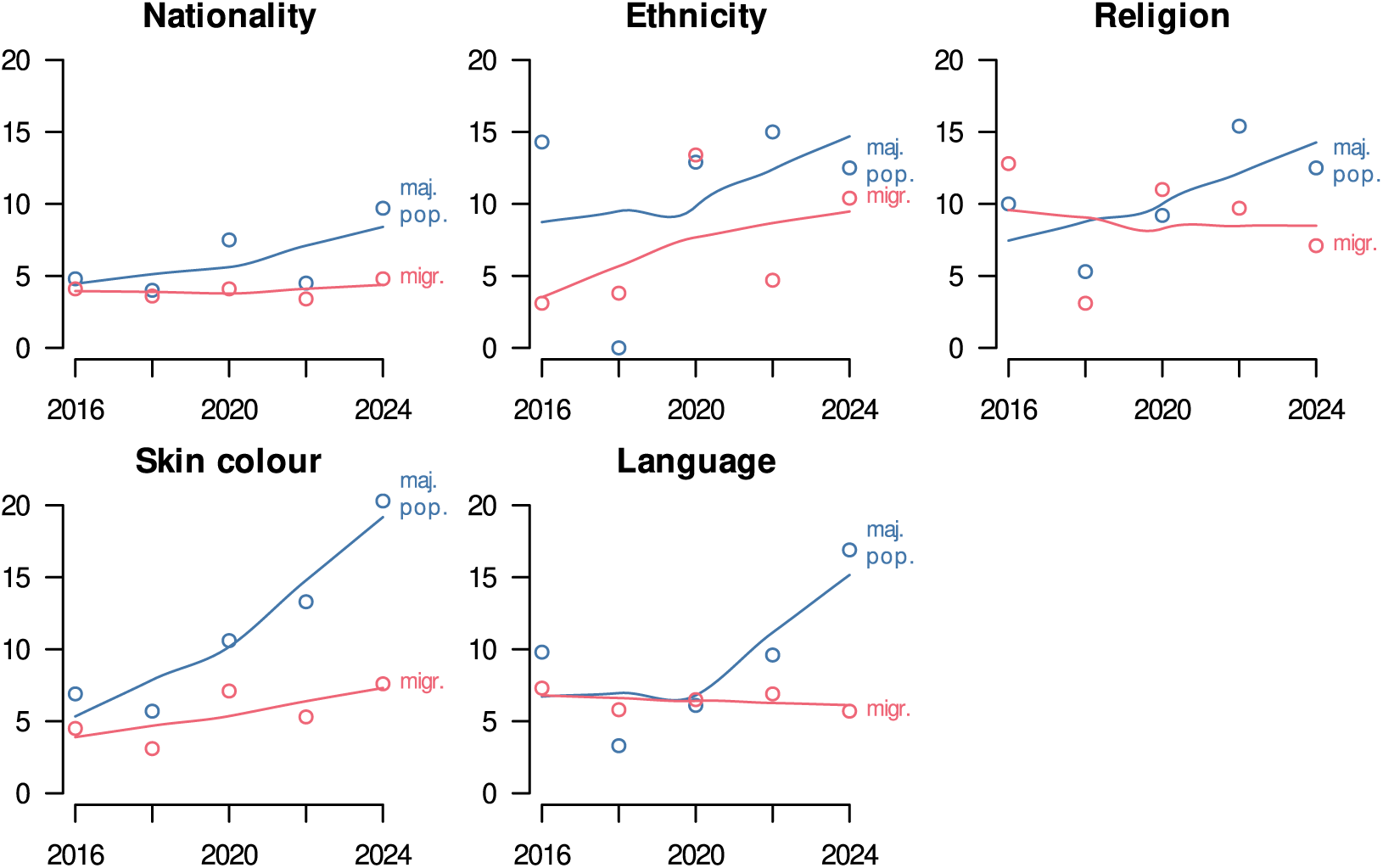
Difference in reported discrimination in healthcare by migration background (2 categories), in percentage points, Switzerland, 2016 to 2024. Different motives of discrimination are used in each panel.

### Country of birth

In this section, we look at the country of birth as an indicator of the migration-related population (Figure 4). The developments over time resemble that shown for migration background in the preceding section. A difference exists in that nationality on its own is not associated with a difference between the migration-related population and the majority population.

**Figure 4:**
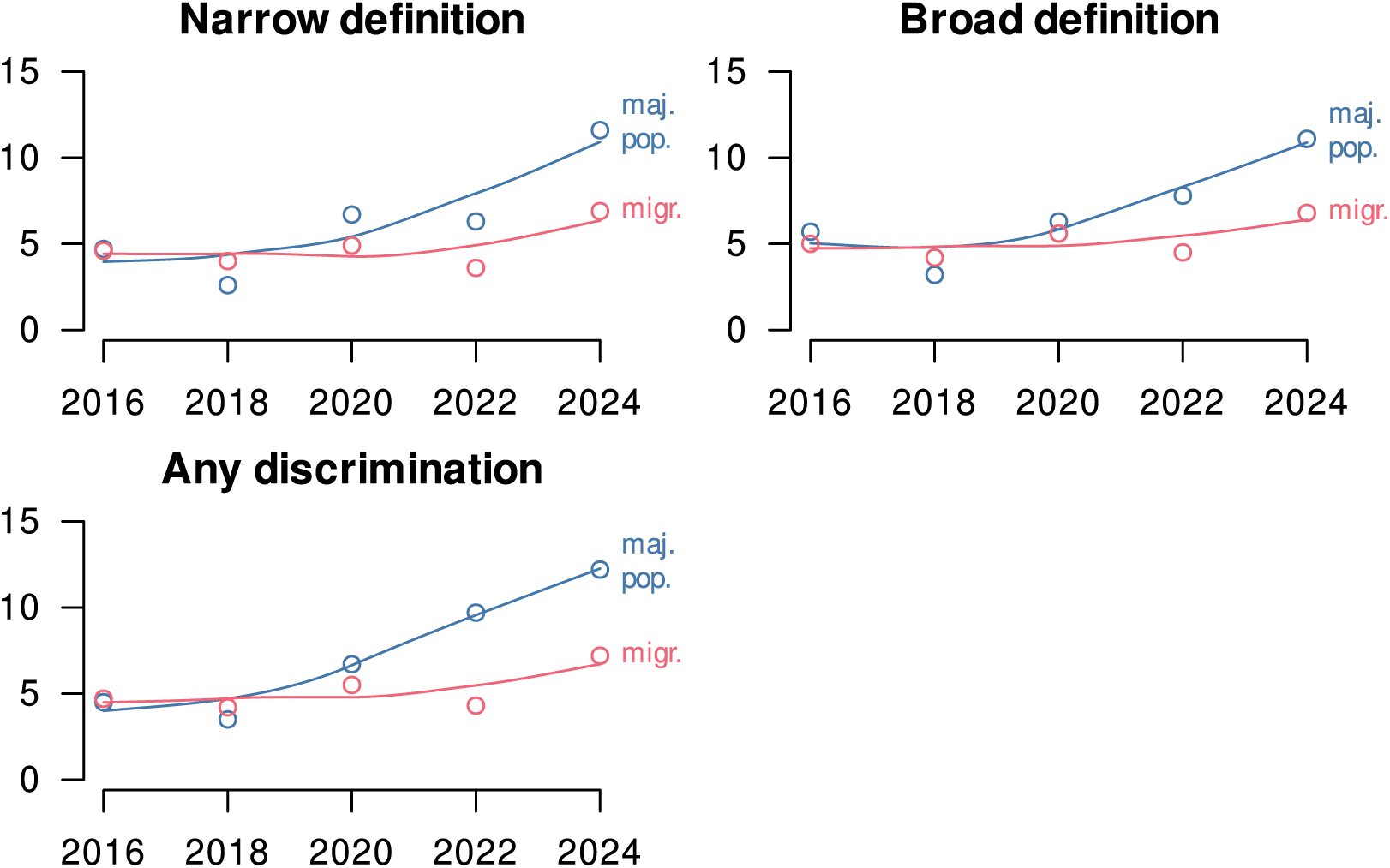
Difference in reported discrimination in healthcare by migration country of birth (Switzerland in blue, abroad in red), in percentage points, Switzerland, 2016 to 2024. Different definitions of discrimination are used in each panel.

**Figure 5:**
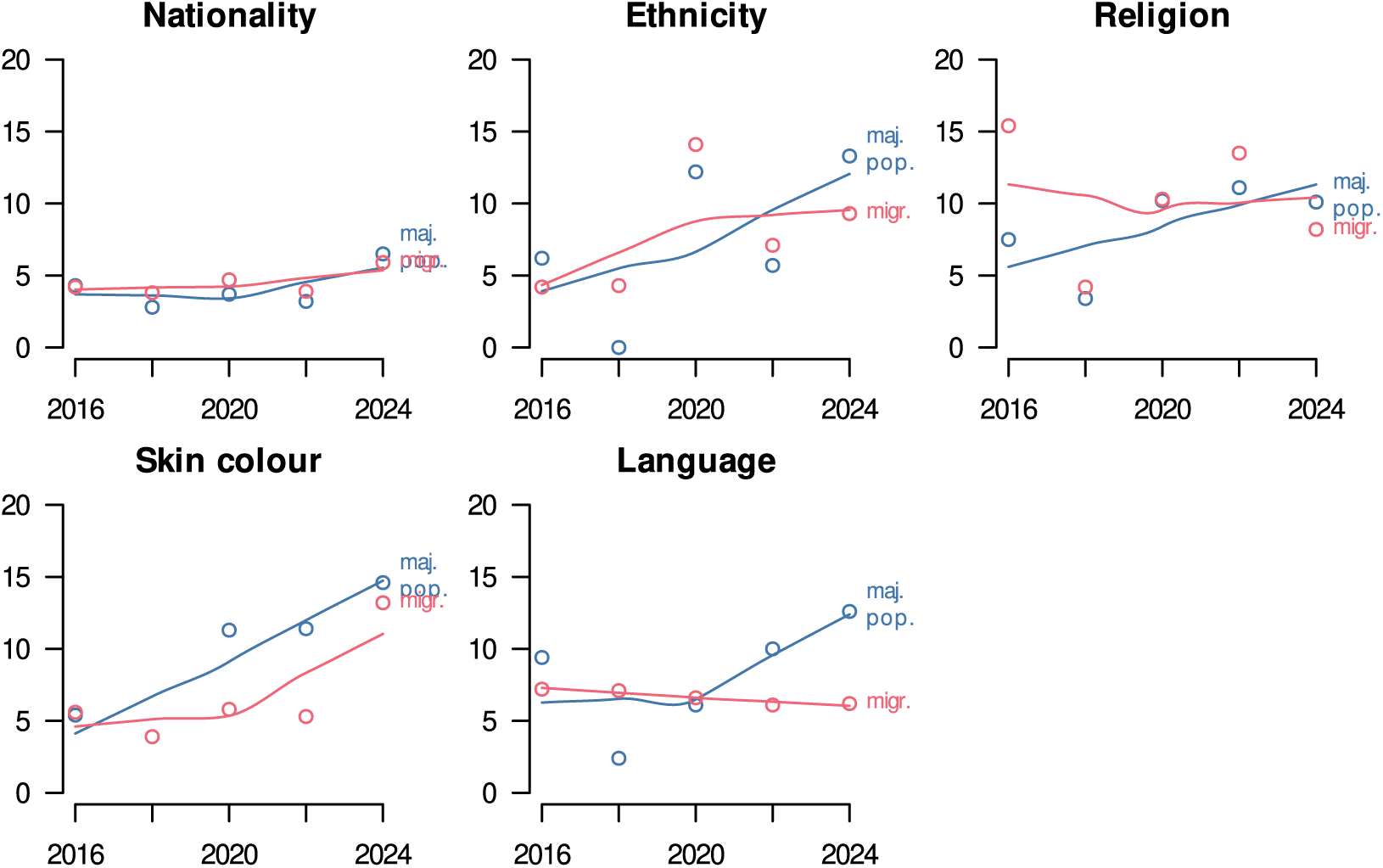
Difference in reported discrimination in healthcare by migration background (2 categories), in percentage points, Switzerland, 2016 to 2024. Different motives of discrimination are used in each panel.

### Swiss Nationality

In Figure 6, we compared foreign citizens (migration-related population) to Swiss citizens. The changes resemble those in the preceding sections.

**Figure 6:**
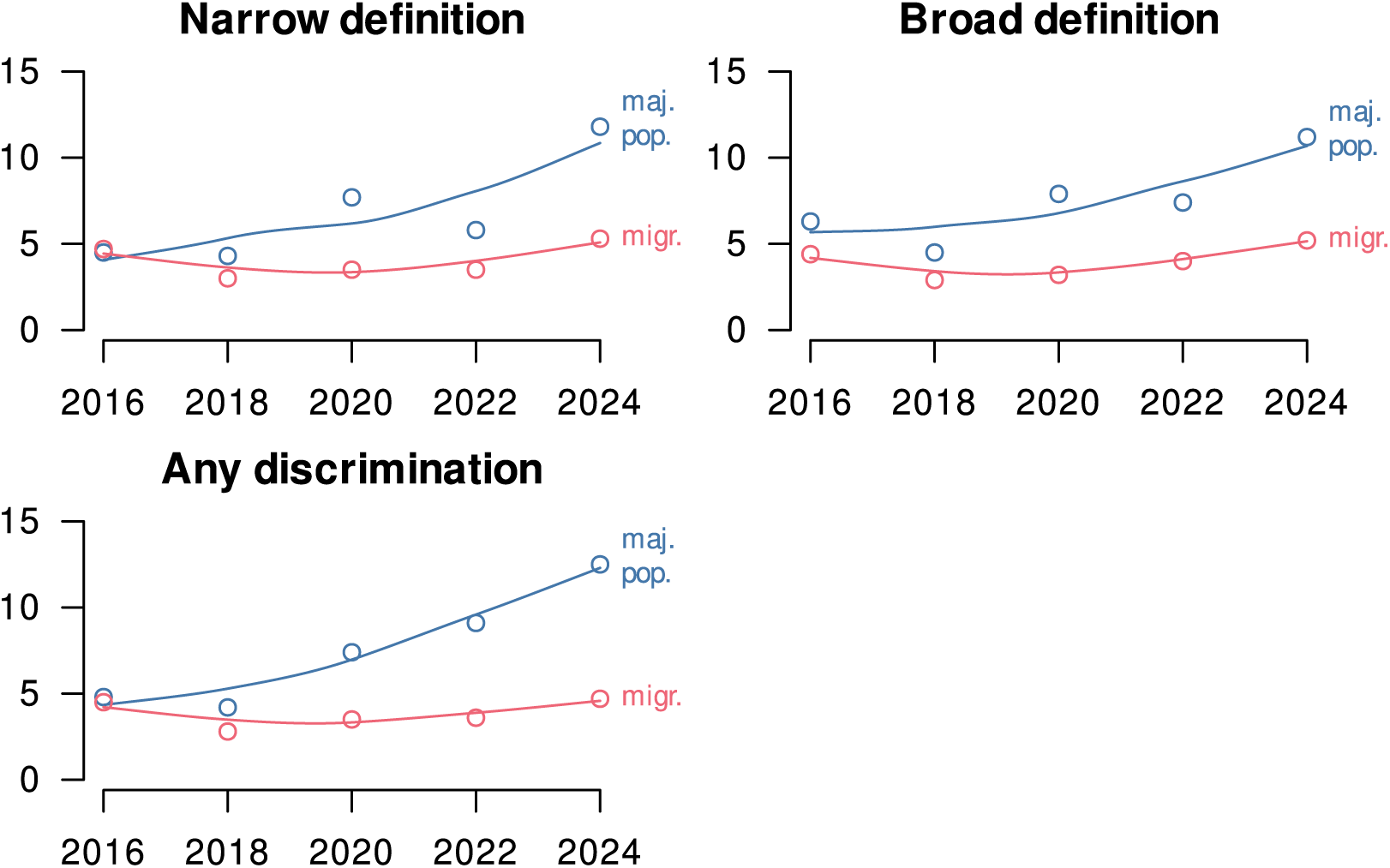
Difference in reported discrimination in healthcare by country of citizenship (nationality), in percentage points, Switzerland, 2016 to 2024. Different definitions of discrimination are used in each panel.

**Figure 7:**
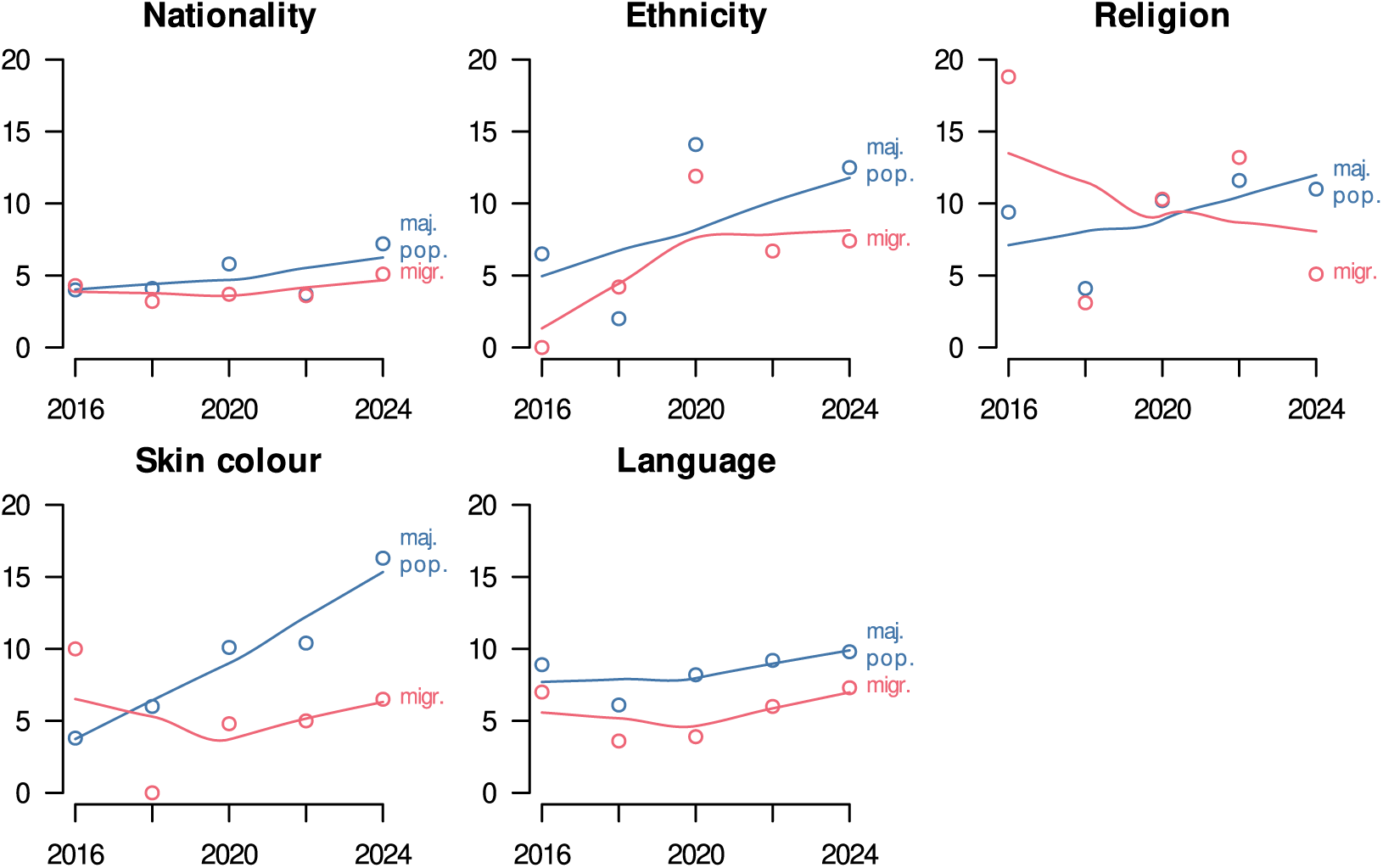
Difference in reported discrimination in healthcare by migration background (2 categories), in percentage points, Switzerland, 2016 to 2024. Different motives of discrimination are used in each panel.

### Migration background (3 categories)

In Figure 8, migration background is divided into migration background of the first generation and migration background of the second generation. These developments illustrate that the relevant change is found in the majority population and not in the migration-related population. The developments for people with a migration background of the first generation and people with a migration background of the second generation is very similar.

**Figure 8:**
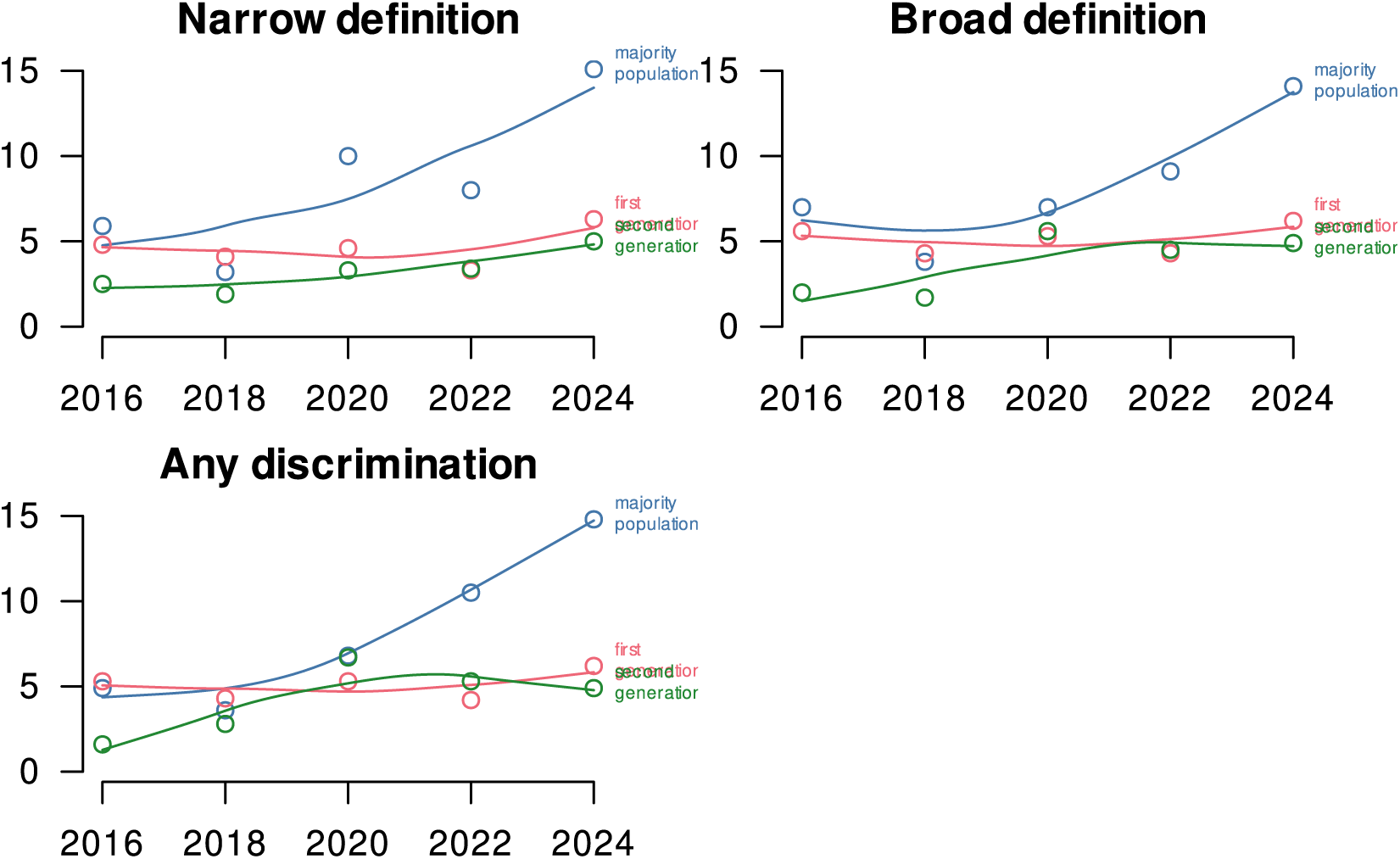
Difference in reported discrimination in healthcare by migration background (3 categories, differentiating 1st and 2nd generation), in percentage points, Switzerland, 2016 to 2024. Different definitions of discrimination are used in each panel.

**Figure 9:**
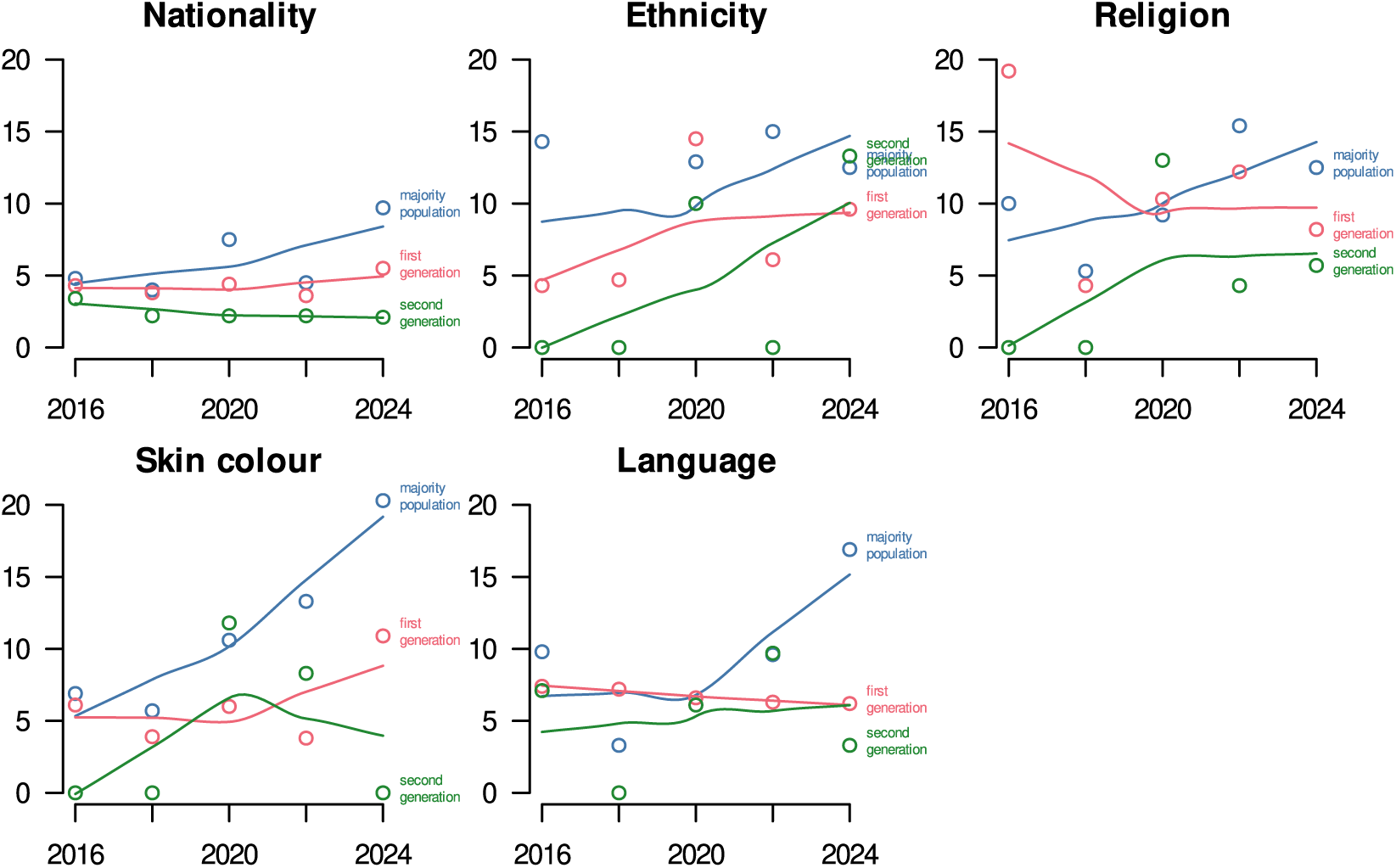
Difference in reported discrimination in healthcare by migration background (2 categories), in percentage points, Switzerland, 2016 to 2024. Different motives of discrimination are used in each panel.

## 5. Multiple regression model for 2024

**Figure 10:**
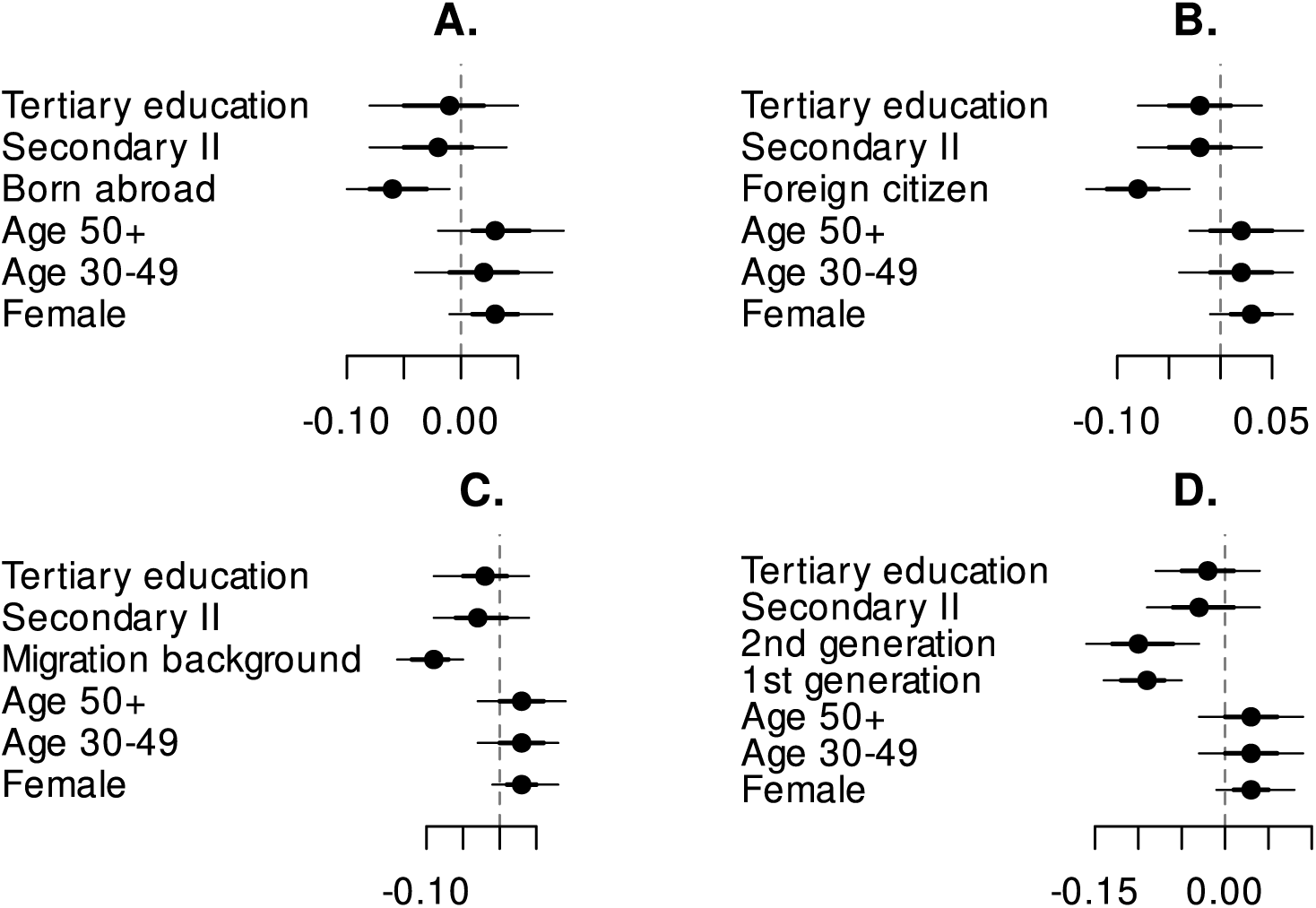
Reported discrimination in healthcare, regression model with 95% credible intervals, each panel with a different approach to defining the migration-related population: born abroad (Panel A), foreign citizenship (B), migration background (C), first and second generation (D), Switzerland, 2024.

## 6. Regression model with combined sample 2016–2024

**Figure 11:**
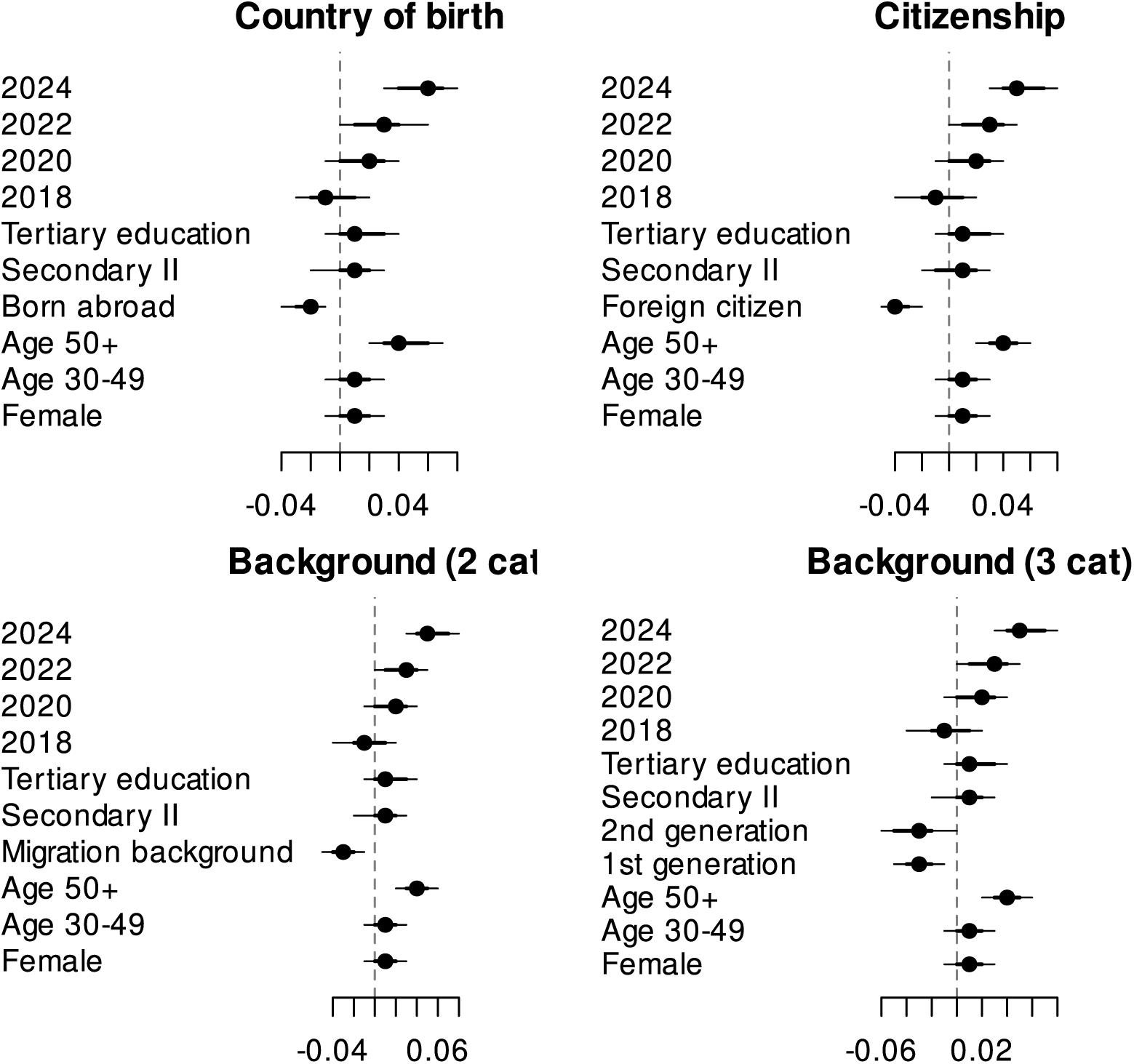
Regression models, with reported discrimination in healthcare as the outcome, Switzerland, 2016 to 2024 combined. Different definitions of the migration-related population are used in each panel.

## 7. Hierarchical model

Here we use hierarchical regression models with individuals nested in years. The results correspond to those of other analytical strategies.

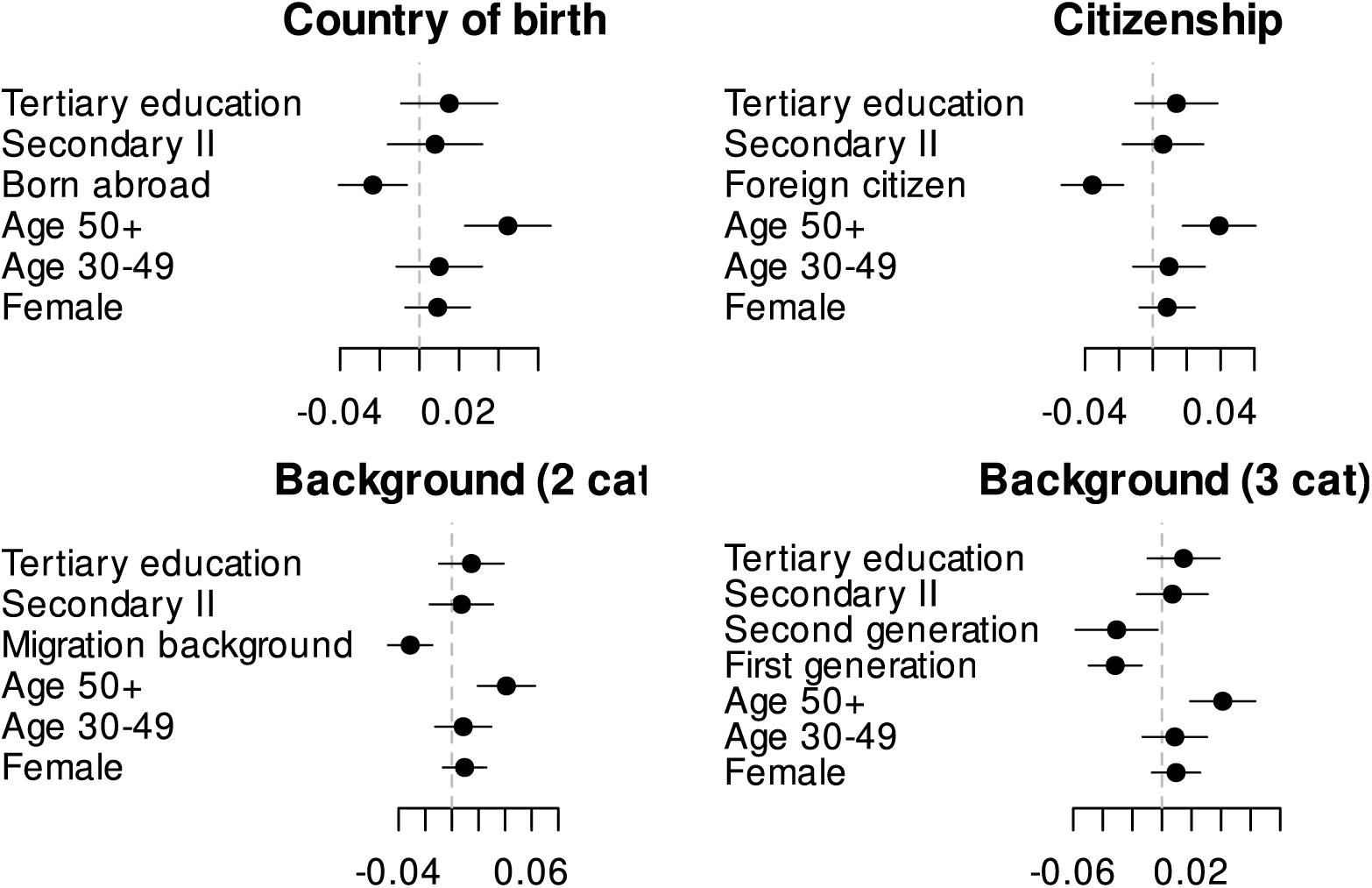

## 8. Accounting for time in Switzerland

The models in this section add the time spent in Switzerland as an additional control variable Figure 12. By definition, this model cannot be determined for respondents by country of birth (no variance within the migration-related population) and for the 3-categorical migration background, respondents of the second generation are excluded (no variation, by definition).

**Figure 12:**
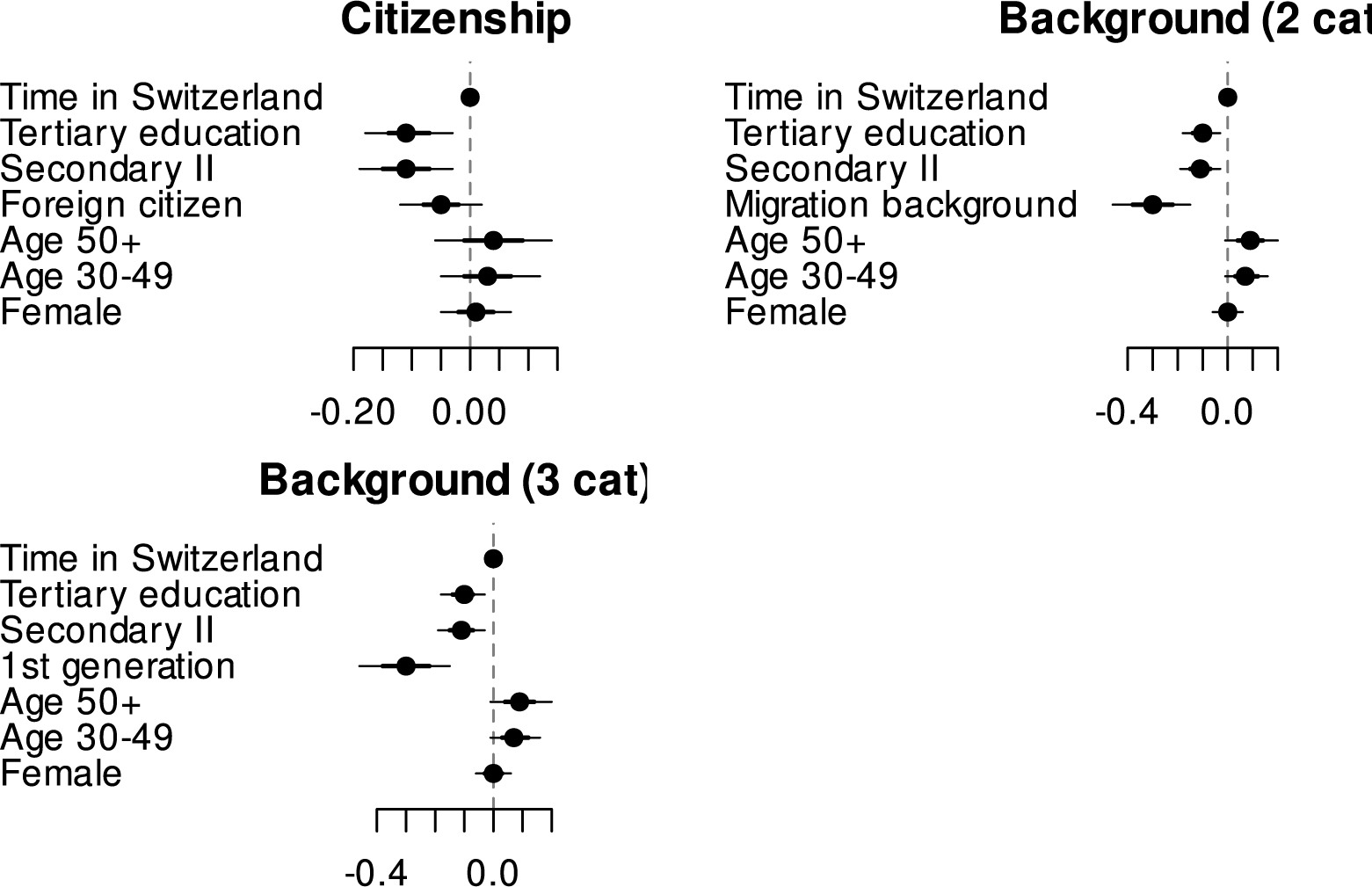
Accounting for time in Switzerland in reported discrimination in healthcare, regression model with 95% credible intervals, Switzerland, 2024. Different definitions of the migration-related population are used in each panel.

## 9. Language regions

We might expect that people in the German-speaking area react more if reported discrimination is a reflection to a (perceived) need to speak standard language with healthcare providers (rather than dialect). We observe no substantive differences in the reported discrimination in the survey Figure 13. There are no substantial differences between language regions.

**Figure 13:**
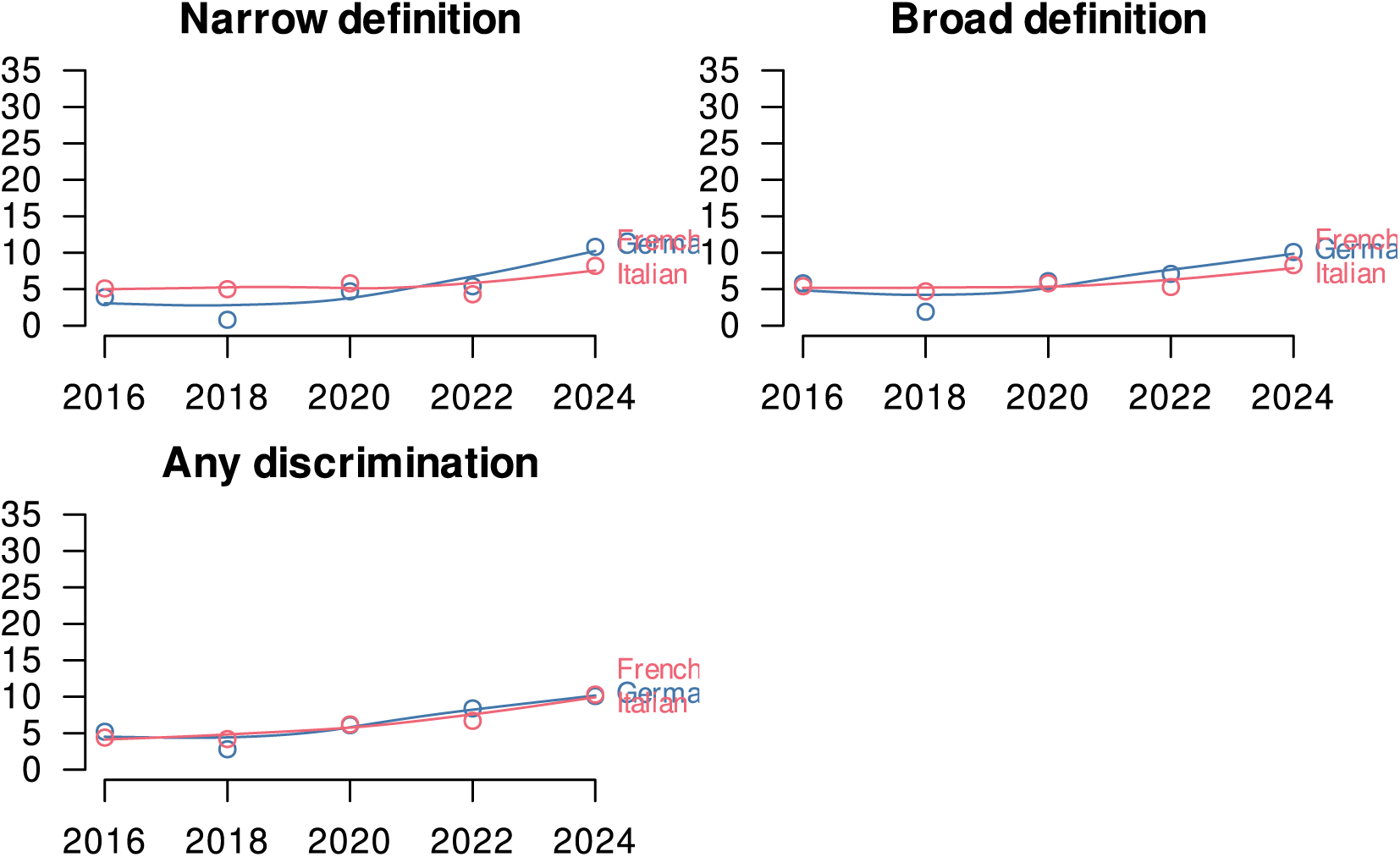
Difference in reported discrimination in healthcare by language region, in percentage points, Switzerland, 2016 to 2024. Different definitions of discrimination are used in each panel.

## 10. Interaction between year and migration−related population

The regression models in Figure 14 develop the models in Figure 11. We look at the interaction between time and migration-related population. There are no new insights, substantially speaking, with the interaction terms picking up the growing levels of reported discrimination for the migration-related population.

**Figure 14:**
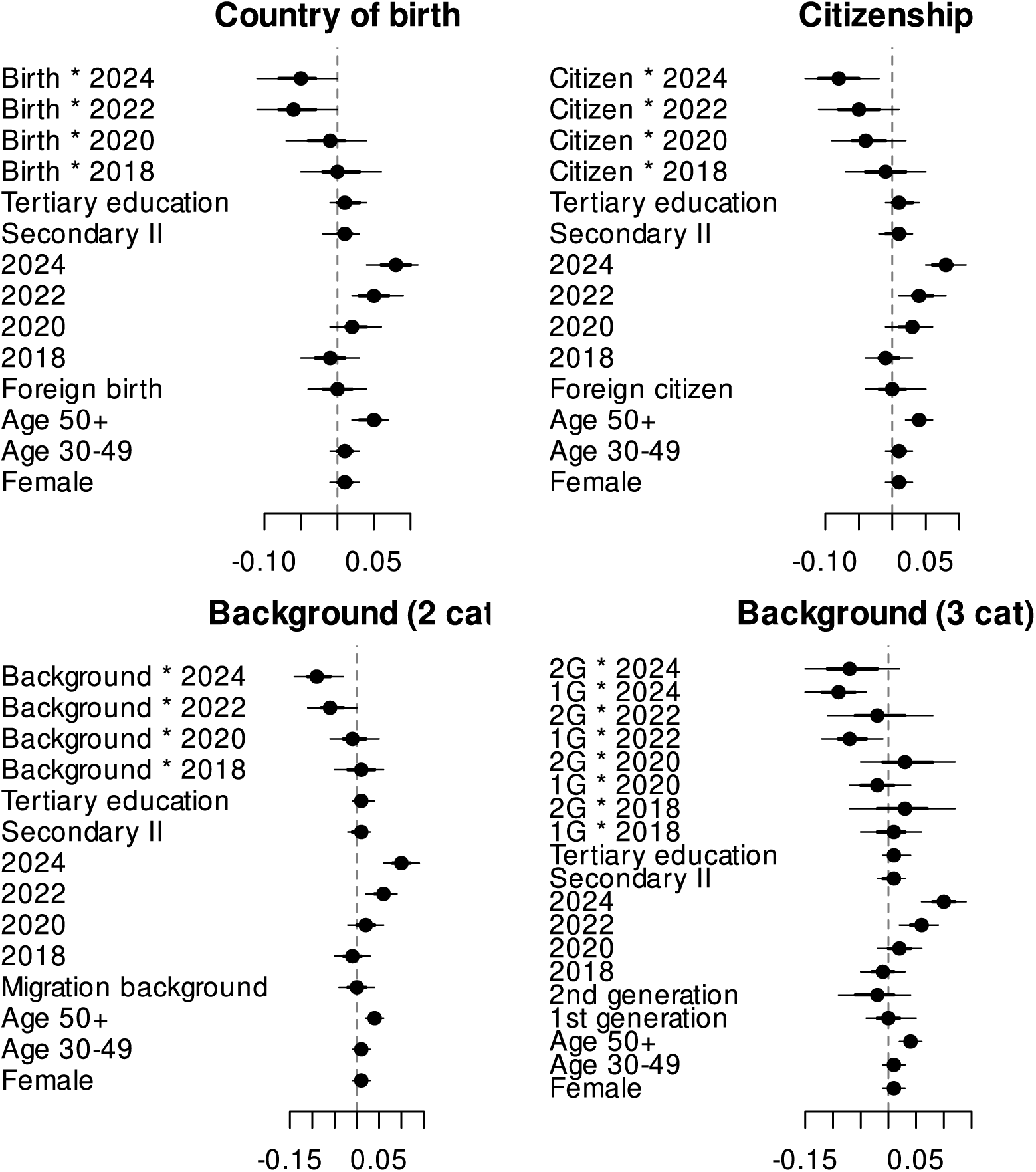
Regression model with interaction effect, Switzerland. 1G = migration background of the first generation, 2G = migration background of the second generation.

## 11. Accounting for ideology (left–right)

**Figure 15:**
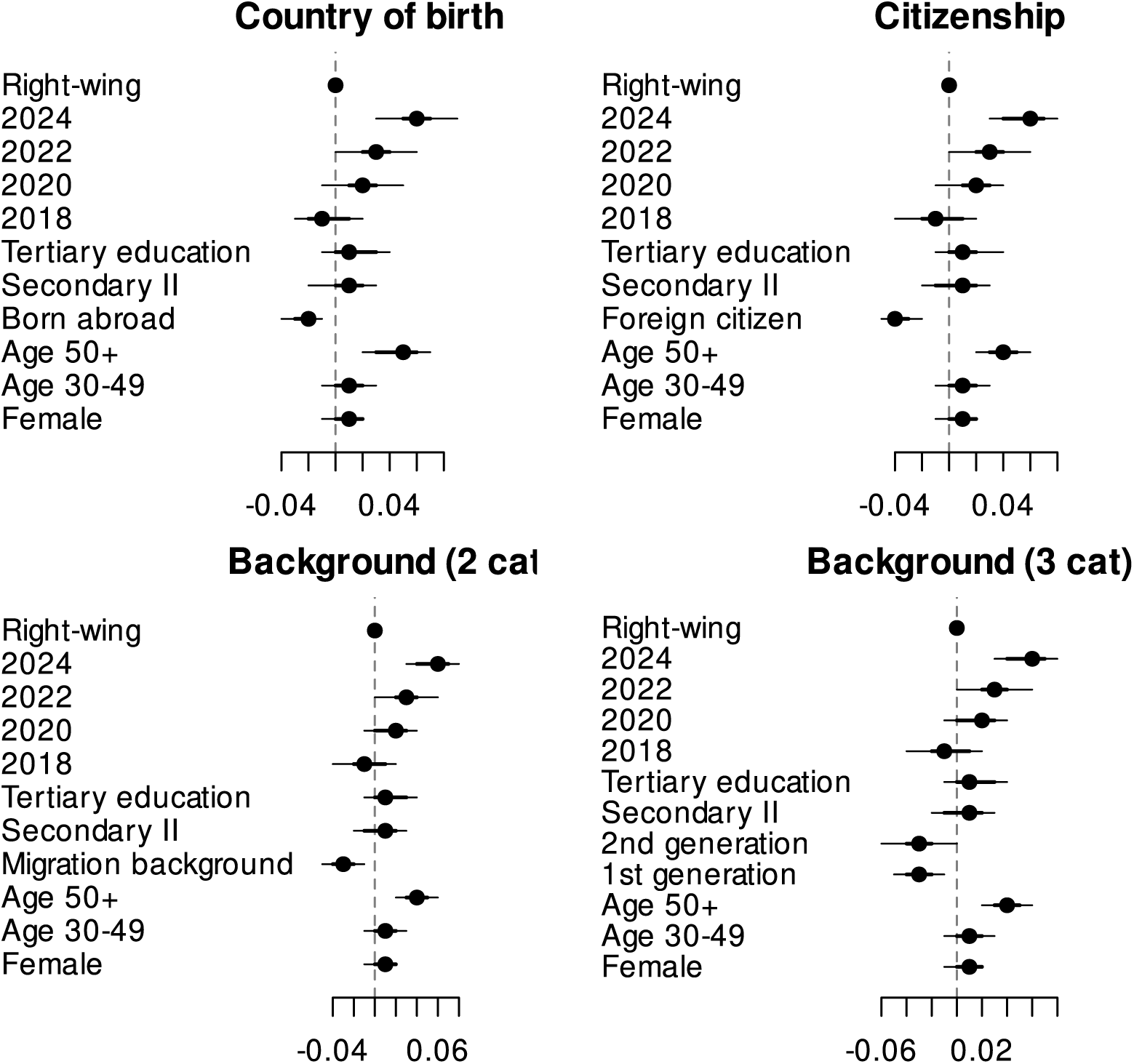
Regression models, with reported discrimination in healthcare as the outcome, Switzerland, 2016 to 2024 combined, controlling for political ideology: right [left]. Different definitions of the migration-related population are used in each panel.

## 12. Regression models for work and housing, 2024

In Figure 16 and Figure 17, we show that the association between migration-related population and reported discrimination is positive for discrimination at work and in housing, contrary to what we find for healthcare.

**Figure 16:**
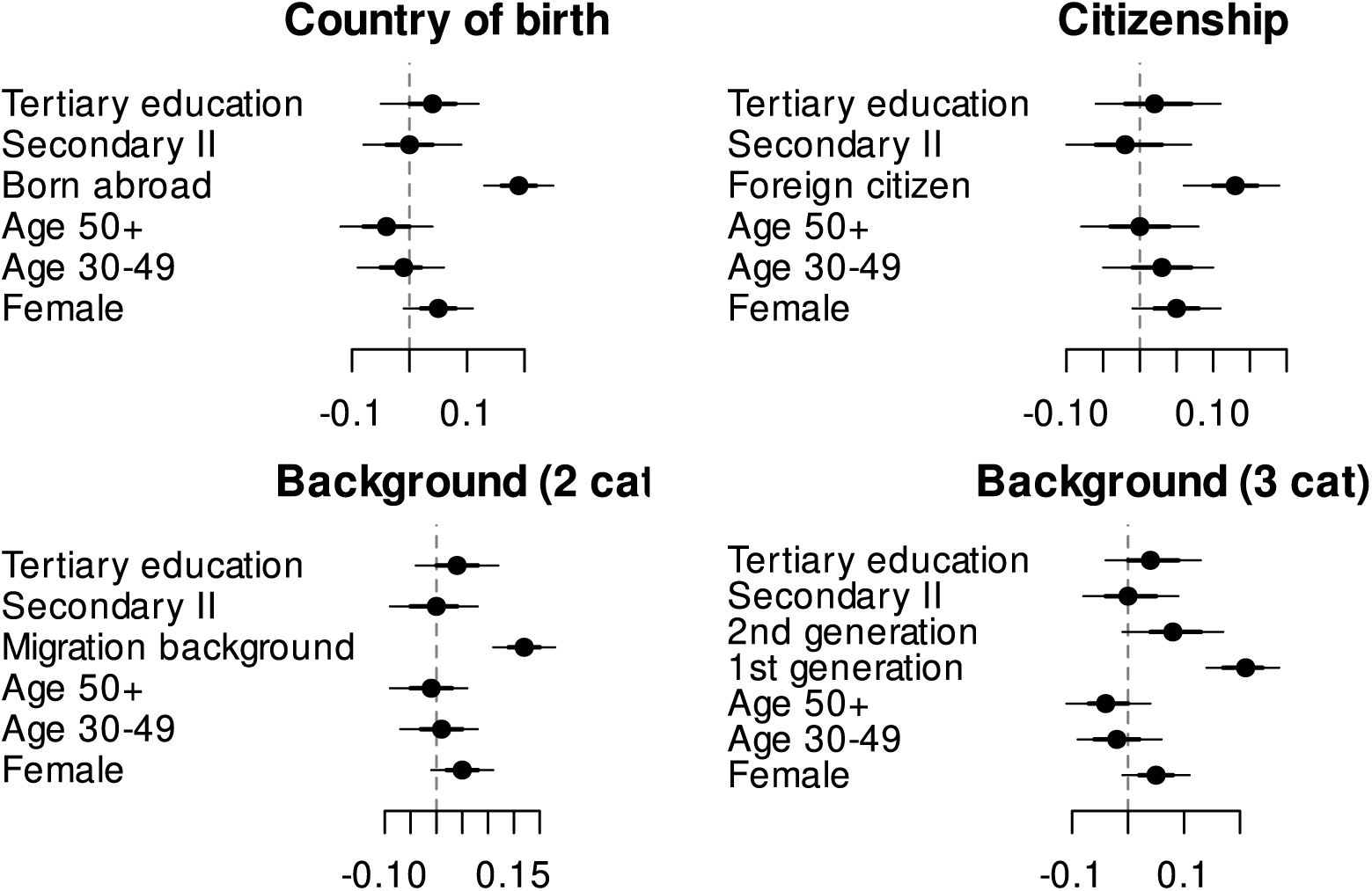
Reported discrimination at work, regression model with 95% credible intervals, Switzerland, 2024. Different definitions of the migration-related population are used in each panel.

**Figure 17:**
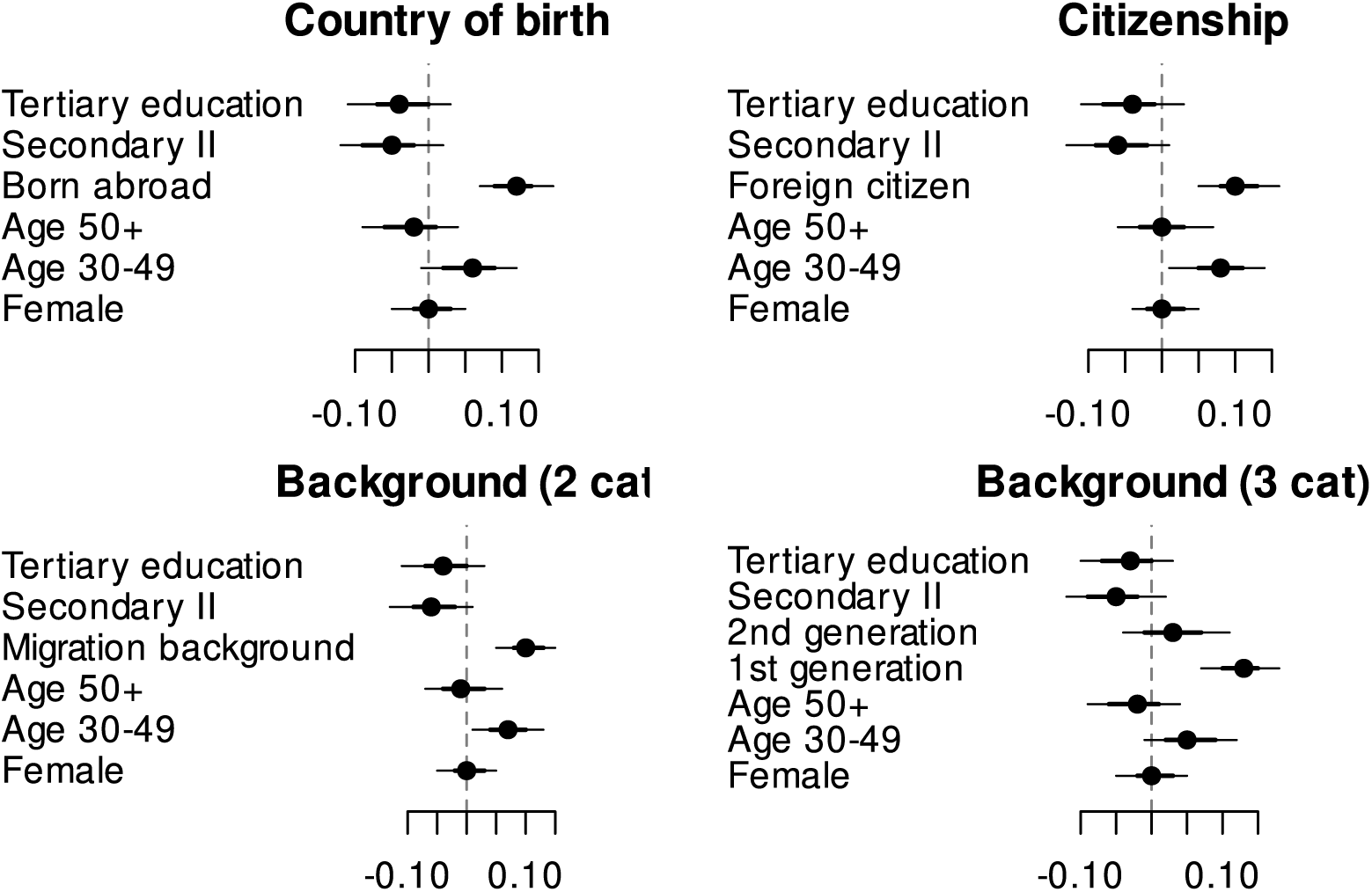
Reported discrimination in housing, regression model with 95% credible intervals, Switzerland, 2024. Different definitions of the migration-related population are used in each panel.

## 13. Changes over time for work and housing

In Figure 18 and Figure 19, we show that the developments over time in Figure 1 are equivalent in different subsamples of varying definitions of ‘racial discrimination’.

**Figure 18:**
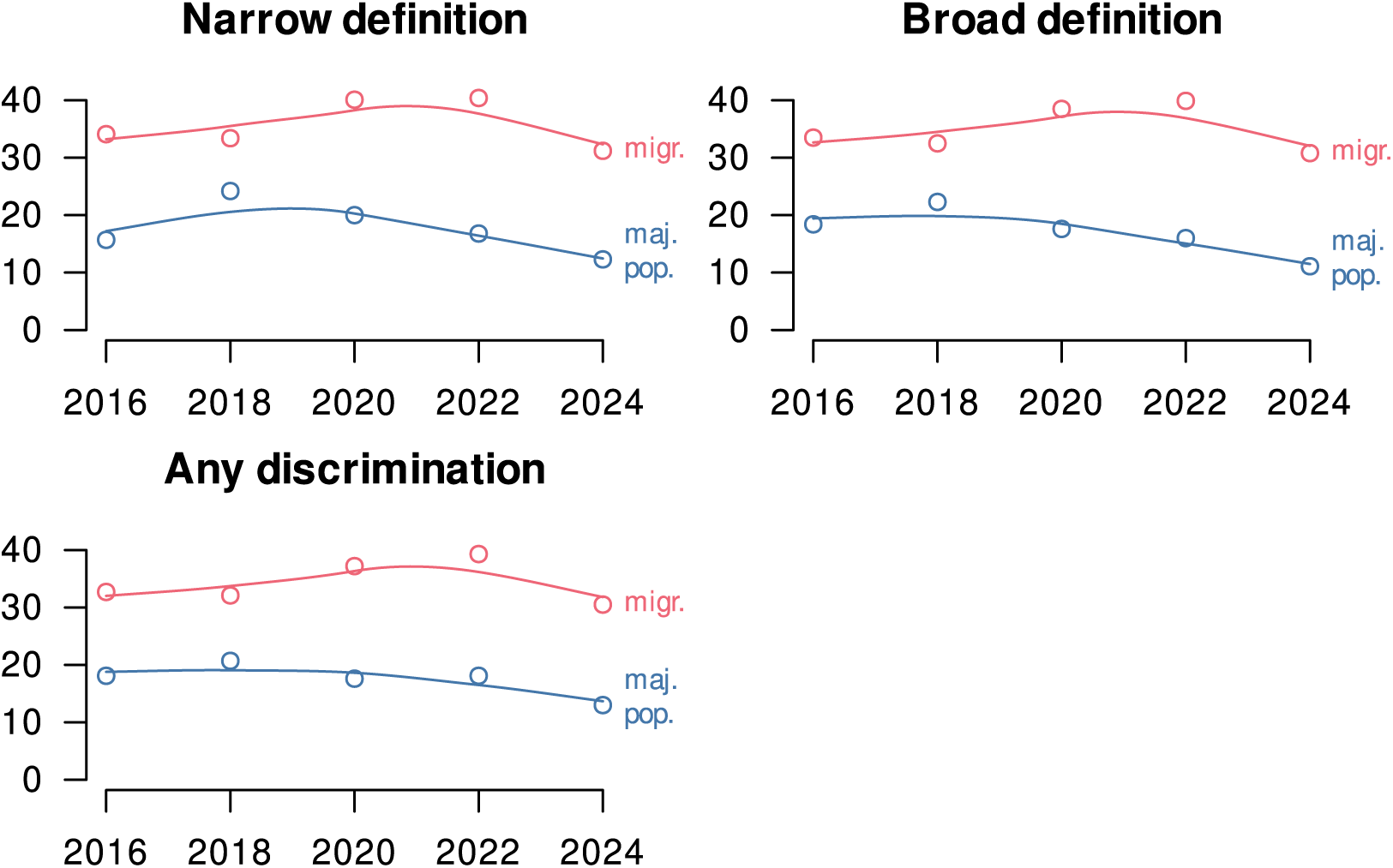
Difference in reported discrimination at work by migration background (2 categories), Switzerland, 2016 to 2024. Different definitions of discrimination are used in each panel.

**Figure 19:**
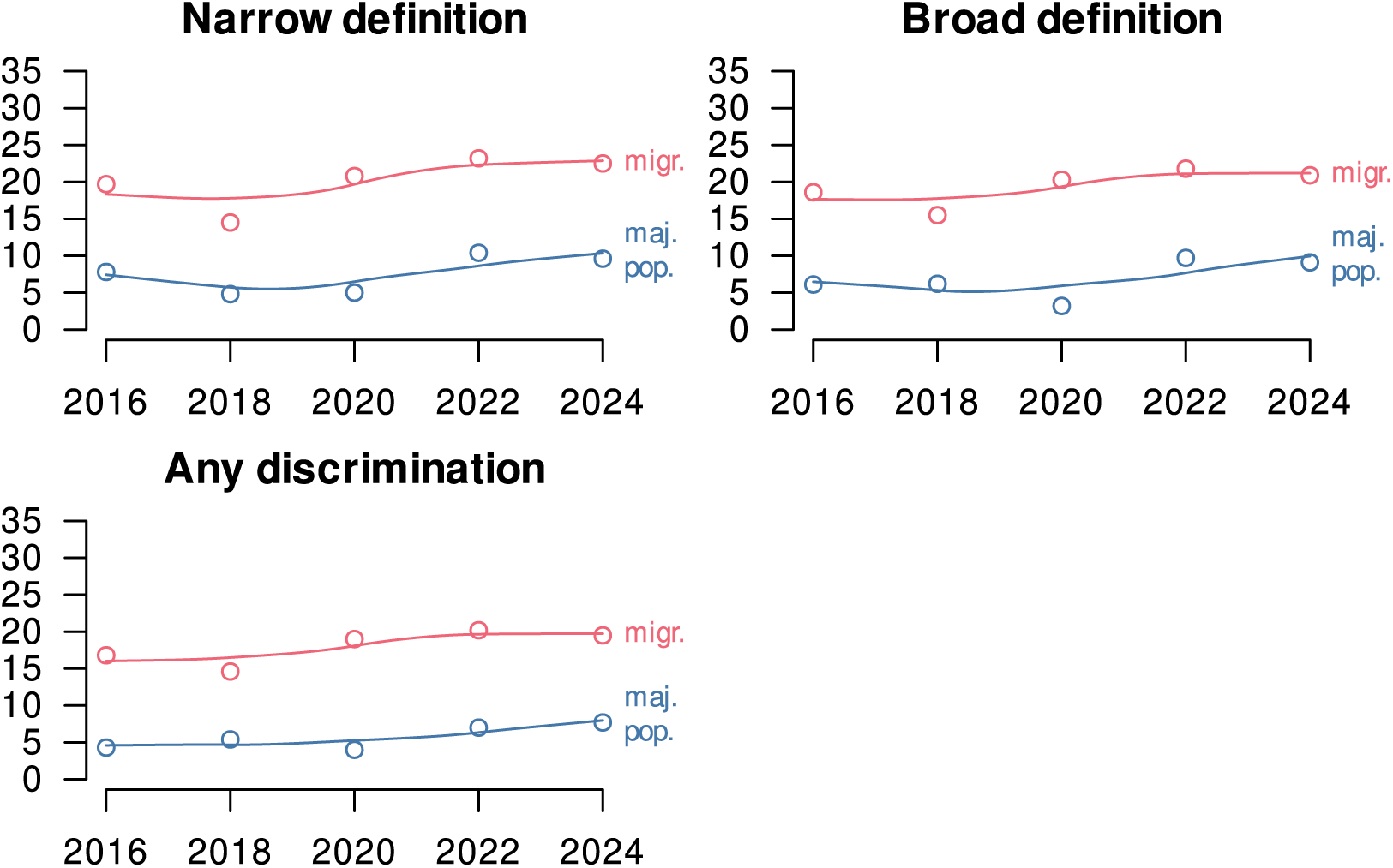
Difference in reported discrimination in housing, by migration background (2 categories), Switzerland, 2016 to 2024. Different definitions of the migration-related population are used in each panel.

## 14. Discrimination in Healthcare in the Migration and Mobility Survey

Here we look at self-reported discrimination in ‘healthcare and care’ (situation) because of ‘racist reasons’, ‘immigrant background, origin, nationality’, or ‘religion’ (motive) reported in the Migration and Mobility Survey (NCCR on the move 2024) in Switzerland 2016 to 2024. The sample consists of people who have moved to Switzerland within 10 years before the survey, and excludes asylum seekers.

**Table 4:** Self-reported discrimination among recent immigrants in Switzerland.

| Year | 2016 | 2018 | 2020 | 2022 | 2024 |
| --- | --- | --- | --- | --- | --- |
| Percentage racially discriminated in healthcare | 2.0 | 2.0 | 2.0 | 2.0 | 2.1 |

